# Mapping a Pandemic: SARS-CoV-2 Seropositivity in the United States

**DOI:** 10.1101/2021.01.27.21250570

**Authors:** Heather Kalish, Carleen Klumpp-Thomas, Sally Hunsberger, Holly Ann Baus, Michael P Fay, Nalyn Siripong, Jing Wang, Jennifer Hicks, Jennifer Mehalko, Jameson Travers, Matthew Drew, Kyle Pauly, Jacquelyn Spathies, Tran Ngo, Kenneth M. Adusei, Maria Karkanitsa, Jennifer A Croker, Yan Li, Barry I. Graubard, Lindsay Czajkowski, Olivia Belliveau, Cheryl Chairez, Kelly Snead, Peter Frank, Anandakumar Shunmugavel, Alison Han, Luca T. Giurgea, Luz Angela Rosas, Rachel Bean, Rani Athota, Adriana Cervantes-Medina, Monica Gouzoulis, Brittany Heffelfinger, Shannon Valenti, Rocco Caldararo, Michelle M. Kolberg, Andrew Kelly, Reid Simon, Saifullah Shafiq, Vanessa Wall, Susan Reed, Eric W Ford, Ravi Lokwani, John-Paul Denson, Simon Messing, Sam G. Michael, William Gillette, Robert P. Kimberly, Steven E. Reis, Matthew D. Hall, Dominic Esposito, Matthew J. Memoli, Kaitlyn Sadtler

## Abstract

Asymptomatic SARS-CoV-2 infection and delayed implementation of diagnostics have led to poorly defined viral prevalence rates. To address this, we analyzed seropositivity in US adults who have not previously been diagnosed with COVID-19. Individuals with characteristics that reflect the US population (*n* = 11,382) and who had not previously been diagnosed with COVID-19 were selected by quota sampling from 241,424 volunteers (ClinicalTrials.gov NCT04334954). Enrolled participants provided medical, geographic, demographic, and socioeconomic information and 9,028 blood samples. The majority (88.7%) of samples were collected between May 10^th^ and July 31^st^, 2020. Samples were analyzed via ELISA for anti-Spike and anti-RBD antibodies. Estimation of seroprevalence was performed by using a weighted analysis to reflect the US population. We detected an undiagnosed seropositivity rate of 4.6% (95% CI: 2.6 – 6.5%). There was distinct regional variability, with heightened seropositivity in locations of early outbreaks. Subgroup analysis demonstrated that the highest estimated undiagnosed seropositivity within groups was detected in younger participants (ages 18-45, 5.9%), females (5.5%), Black/African American (14.2%), Hispanic (6.1%), and Urban residents (5.3%), and lower undiagnosed seropositivity in those with chronic diseases. During the first wave of infection over the spring/summer of 2020 an estimate of 4.6% of adults had a prior undiagnosed SARS-CoV-2 infection. These data indicate that there were 4.8 (95% CI: 2.8-6.8) undiagnosed cases for every diagnosed case of COVID-19 during this same time period in the United States, and an estimated 16.8 million undiagnosed cases by mid-July 2020.

## Introduction

COVID-19, the disease caused by SARS-CoV-2 infection, presents with a spectrum of illness ranging from asymptomatic to severe disease and death. As with most respiratory viral diseases, it is difficult to estimate the true prevalence of the disease during a pandemic and the extent of its spread is only known after extensive study^1-3^. The majority of patients infected develop robust antibody responses against the viral spike (S), nucleocapsid (N), and envelope (E) proteins that can be detected via serologic testing^4-8^. Anti-S antibodies persist for months, and can neutralize infection^9^. Frequently, these neutralizing antibodies bind the receptor binding domain (RBD) of the spike protein, but antibodies against the spike S2 domain have also been observed^10-15^.

To characterize the spread of SARS-CoV-2 infection in the United States, we evaluated seropositivity in a national survey of participants who had not previously been diagnosed with SARS-CoV-2 infection. We used quota sampling from a large pool of volunteers to obtain a representative sample and performed statistical weighting to generate prevalence estimates which provide a clear picture of the extent of SARS-CoV-2 infection. To ensure accurate classification of seropositivity, we utilized our dual-antigen ELISA protocol that evaluated IgG and IgM antibodies against both the full spike ectodomain and the RBD^7,16^. These foundational considerations generated critical data needed to estimate spread during the pandemic and gain insight into the potential future outcomes.

These results, including the subgroup analysis, give us a previously undescribed view into the spread of the pandemic by more clearly identifying the large numbers of individuals with undiagnosed infections during the initial months of the pandemic. These data are of great importance as we consider the impact vaccination may have on the future course of the pandemic and plan for current and future available vaccines to be administered. In addition, these data can also help us better assess the public health measures taken during the pandemic and how to take the best approaches forward to any future public health emergencies.

## METHODS

### Study Protocol

This study was designed to determine the seroprevalence of anti-SARS-CoV-2 antibodies in adults 18 years of age or older in the United States who had not been previously diagnosed with COVID-19. The primary endpoint was the weighted estimate of seroprevalence in the US. Secondary endpoints were weighted estimates for subgroups categorized by demographics/risk factors. An initial period enrolled a convenience sample of 593 volunteers prior to the quota sample. Participants across the US (all 50 states and DC) were then enrolled via telephone consent from a pool of volunteers who provided basic demographic data in response to the study announcement. Recruitment calls were made from three sites: NIAID Laboratory of Infectious Diseases Clinical Studies Unit, the University of Pittsburgh CTSI, and the University of Alabama at Birmingham CCTS. Selection of participants is described below. Selected participants were contacted by the study team, consented, and sent a blood microsampling kit and online questionnaire in REDCap (project-redcap.org). For a small subset of participants (*n* = 214) working on the NIH campus, serum was collected via venipuncture. This study (ClinicalTrials.gov NCT04334954) was approved by the National Institutes of Health Institutional Review Board and conducted in accordance with the provisions of the Declaration of Helsinki and Good Clinical Practice guidelines. All participants provided verbal informed consent prior to enrollment.

### Participant Selection

All volunteers were emailed an initial survey to collect basic demographic characteristics. Survey responses were de-identified and aggregated by sub-category of state, type of locality approximated from zip codes, age, sex, race, and ethnicity (**Figure 1**). Target sample sizes for these sub-categories were determined from the U.S. census, and were updated every evening based on the characteristics of people who had already enrolled to assure that individuals in each sub-category were enrolled evenly over time. Within each sub-category, participants were initially assigned a selection probability calculated from the target number as a proportion of the available pool. Specific sub-categories that had insufficient numbers were aggregated to estimate their impact on the overall distribution of the 6 main characteristics. If a particular characteristic had insufficient numbers, sample probabilities were boosted for volunteers who had the characteristic. For each day’s call list, the most representative of 20,000 randomly generated lists was used, each list drawn without replacement from the volunteer pool based on the sampling probabilities previously defined. Representativeness was assessed by estimating a weighted sum of squared differences from the desired targets and picking the list with the lowest deviation. Unselected participants were eligible to be called at a later date. This algorithm is designed such that each cohort of invited participants is representative of the diversity of the US population with respect to the 6 sampling variables (see Statistical Supplement Section 3.4).

**Figure 1:**
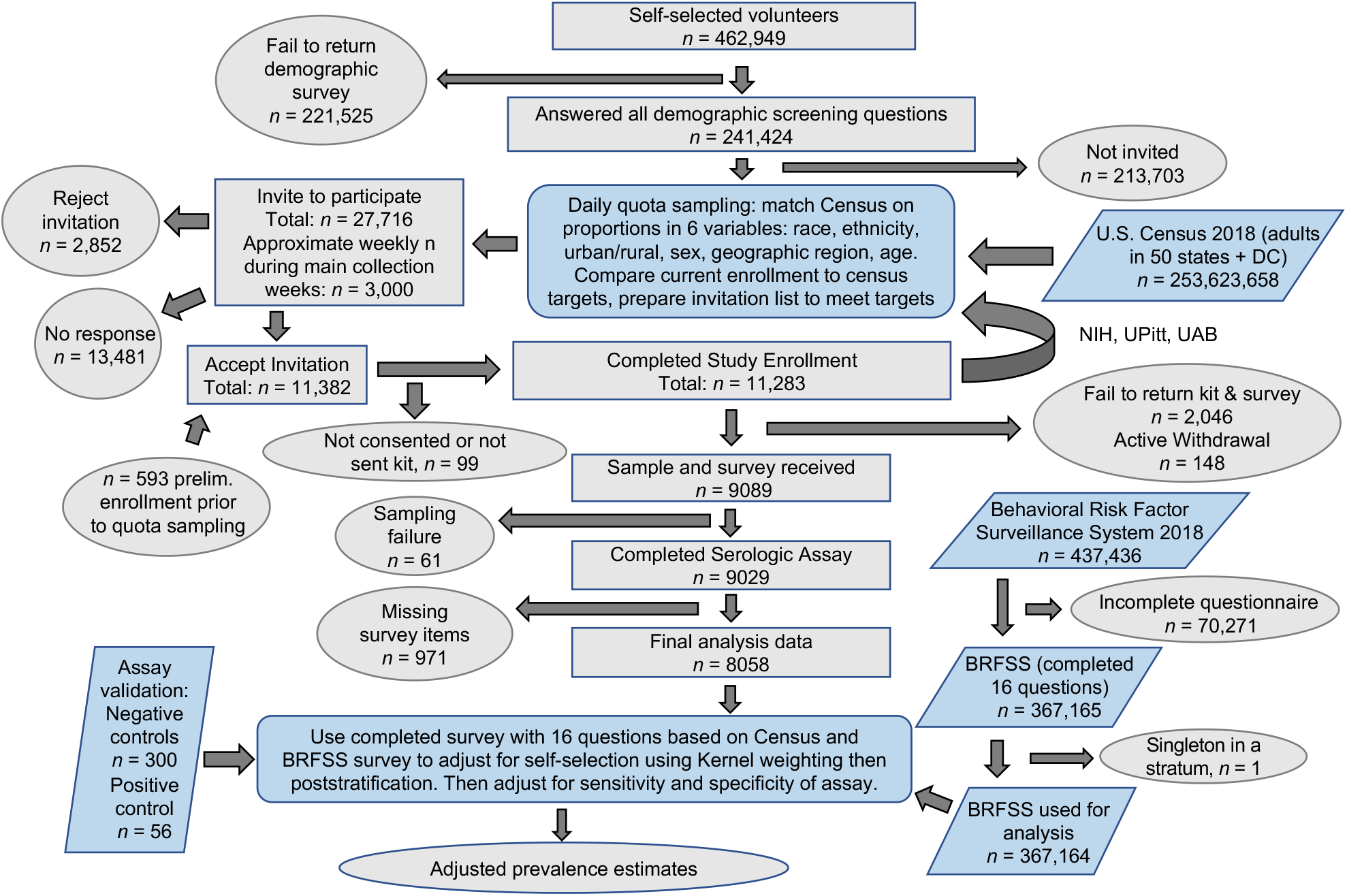
Serosurvey overview and statistical workflow. A flow chart of donor recruitment through data analysis displaying steps in data acquisition and any attrition from data sets if applicable. Key: Ovals = starts and ends, gray rectangles = subsets of participants in this study, blue parallelograms = individuals from outside data sets that contribute to adjusted prevalence estimates, blue rounded rectangles = analysis processes.

### Sample Collection

Participants provided blood samples by mail using a Mitra microsampling kit (Neoteryx, Torrance, CA) or standard venipuncture. Microsampling kits contained visual instructions on the sampling process, bandages, gauze, lancets, and four 20 μl microsampling devices for a total collection of 80 μl of whole blood. Participants utilized the lancet to draw blood from their fingertip and collect blood onto each of the four microsamplers. Participants returned the dried microsamplers with desiccant via overnight shipping. Those who underwent venipuncture did so in the NIH Clinical Center phlebotomy lab where 18 ml of blood was collected in a serum separator and whole blood tube. Once received in the laboratory serum samples were processed, and microsamplers were stored dry at −80°C until elution and analysis.

### Serologic Assays

Antibodies from samples were analyzed using ELISA as previously described^7,16-18^. In order to maintain longitudinal quality control and ensure that the assays remained stable across multiple months of assay implementation, positive and negative controls were included on each assay plate and monitored for stability (**Supplemental Fig. 1**). Seropositivity cut points were defined by evaluating 300 true negative samples and 56 true positive samples. Positivity thresholds were based on the mean optical density (absorbance) plus 3 standard deviations (see Supplemental Materials for details). The final criterion of a Spike^+^ and RBD^+^ for any combination of IgG or IgM gave estimated sensitivity and specificity of 1, with raw values for recombinant antibody results reported in **Supplemental Fig. 2** and **Supplemental Table 1**. Additionally, IgA was evaluated via previously described ELISA to further phenotype the participant’s serologic status.

**Table 1:** Characteristics of serosurvey population in comparison to United States population. Census and Behavioral Risk Factor Surveillance System (BRFSS, 2018) data on selection criteria were utilized for quota-based sampling. Other values from BRFSS were utilized for statistical weighting. The comparisons between the estimated proportions in the United States (BRFSS) versus our sample population for the SARS-CoV-2 serosurvey are displayed in this table.

|  | US Population (BRFSS) |  |  | CoV2 Serosurvey Population |  |
| --- | --- | --- | --- | --- | --- |
|  | n | % | weighted (%) | n | % |
| **Selection Criteria** |  |  |  |  |  |
| Region |  |  |  |  |  |
| North East | 91307 | 21.19 | 17.6 | 1508 | 16.7 |
| Midwest | 67110 | 15.57 | 16.97 | 1445 | 16.01 |
| Mid-Atlantic | 80979 | 18.79 | 16.91 | 1833 | 20.3 |
| South/Central | 60482 | 14.03 | 15.35 | 1293 | 14.32 |
| Mountain/Southwest | 86204 | 20 | 15.89 | 1392 | 15.42 |
| West/Pacific | 44866 | 10.41 | 17.27 | 1557 | 17.25 |
| Age Group |  |  |  |  |  |
| 18 - 45 | 125081 | 28.59 | 46 | 3837 | 42.51 |
| 45 - 70 | 207749 | 47.49 | 39.84 | 3783 | 41.91 |
| 70 - 95 | 104605 | 23.91 | 14.17 | 1407 | 15.59 |
| Sex |  |  |  |  |  |
| Male | 197411 | 45.24 | 48.66 | 4318 | 47.83 |
| Female | 238911 | 54.76 | 51.34 | 4710 | 52.17 |
| Urban/Rural |  |  |  |  |  |
| Urban | 365714 | 84.9 | 93.48 | 8550 | 94.78 |
| Rural | 65234 | 15.1 | 6.52 | 471 | 5.22 |
| Race |  |  |  |  |  |
| White only | 345710 | 81 | 73.41 | 6986 | 77.4 |
| Black only | 37862 | 8.87 | 12.9 | 830 | 9.2 |
| Others | 43219 | 10.13 | 13.69 | 1210 | 13.41 |
| Ethnicity |  |  |  |  |  |
| Hispanic | 36941 | 8.53 | 17.06 | 1495 | 16.56 |
| Not Hispanic | 395931 | 91.47 | 82.94 | 7532 | 83.44 |
| **Additional Weighting Criteria** |  |  |  |  |  |
| Children |  |  |  |  |  |
| Yes | 113408 | 26.21 | 35.81 | 2943 | 32.88 |
| No | 319281 | 73.79 | 64.19 | 6009 | 67.12 |
| Education |  |  |  |  |  |
| <=HS | 151606 | 34.79 | 41.07 | 240 | 2.68 |
| College | 119979 | 27.53 | 30.88 | 1284 | 14.35 |
| >=College | 164229 | 37.68 | 28.05 | 7422 | 82.96 |
| Homeowner |  |  |  |  |  |
| Own | 305545 | 70.36 | 66.49 | 6635 | 74.12 |
| Rent | 107208 | 24.69 | 27.32 | 1861 | 20.79 |
| Others | 21535 | 4.96 | 6.19 | 456 | 5.09 |
| Employment |  |  |  |  |  |
| Employed | 219493 | 50.75 | 57.74 | 6364 | 71.09 |
| NLF | 174920 | 40.45 | 31.38 | 2129 | 23.78 |
| Unemployed | 38053 | 8.8 | 10.88 | 459 | 5.13 |
| Health Insurance |  |  |  |  |  |
| Yes | 400028 | 91.86 | 87.85 | 8697 | 97.31 |
| No | 35433 | 8.14 | 12.15 | 240 | 2.69 |
| Flu Vaccinated |  |  |  |  |  |
| Yes | 234727 | 59 | 50.62 | 6198 | 73.73 |
| No | 163124 | 41 | 49.38 | 2208 | 26.27 |
| Cardiovascular Disease |  |  |  |  |  |
| Yes | 52284 | 12.07 | 9.07 | 354 | 3.98 |
| No | 380985 | 87.93 | 90.93 | 8541 | 96.02 |
| Pulmonary Disease |  |  |  |  |  |
| Yes | 84102 | 19.33 | 18.53 | 1671 | 18.96 |
| No | 350913 | 80.67 | 81.47 | 7140 | 81.04 |
| Immune Disease |  |  |  |  |  |
| Yes | 170115 | 39.14 | 29.29 | 2039 | 23.1 |
| No | 264571 | 60.86 | 70.71 | 6787 | 76.9 |
| Diabetes |  |  |  |  |  |
| Yes | 60703 | 13.9 | 11.41 | 482 | 5.41 |
| No | 375876 | 86.09 | 88.59 | 8430 | 94.59 |

**Figure 2:**
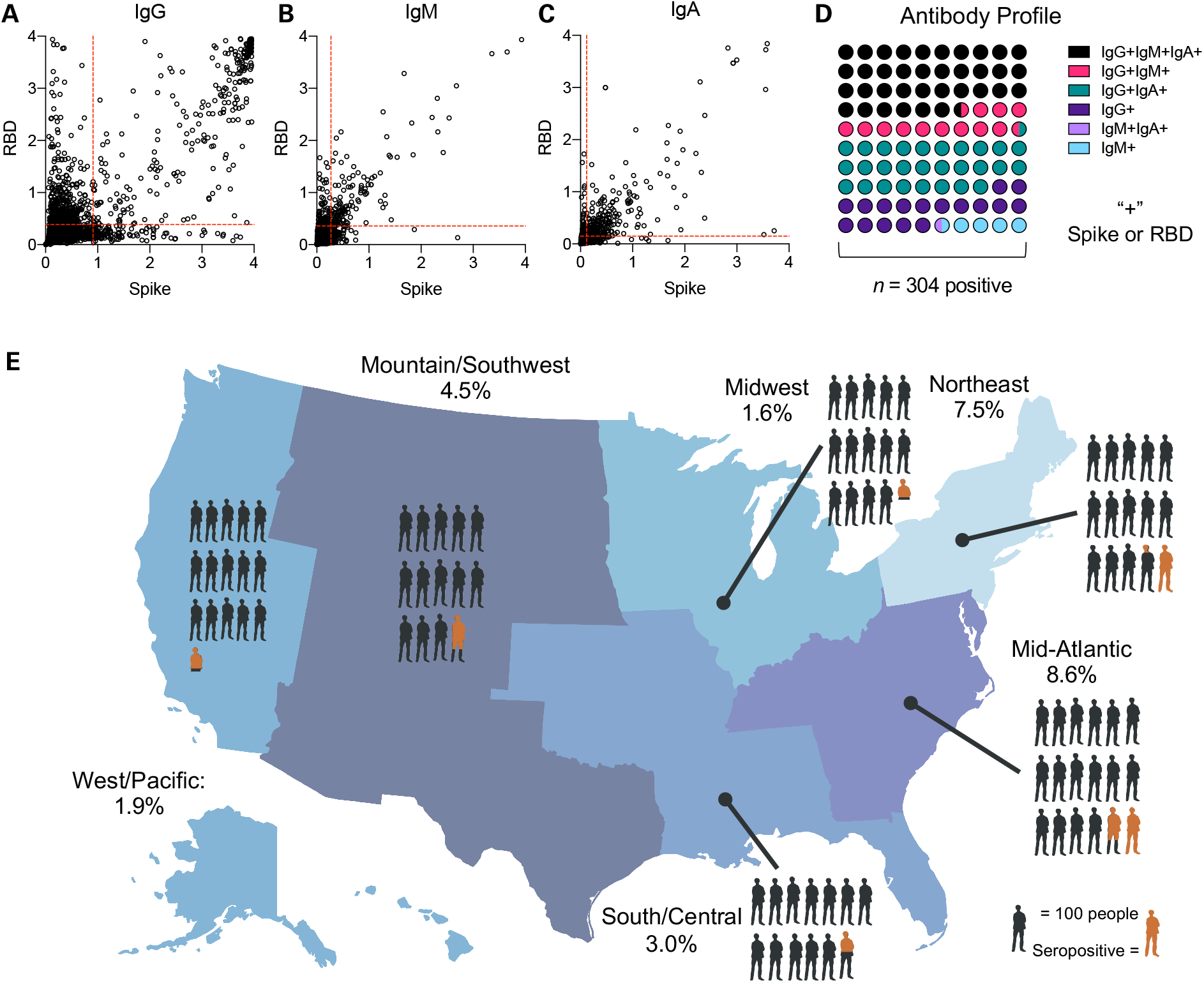
Geographic distribution of undiagnosed seropositivity in the United States in summer 2020. Raw serology data for (a) IgG and (b) IgM and (c) IgA against SARS-CoV-2 Spike and Receptor Binding Domain (RBD). Cut points for positivity are shown as red dashed lines, data are optical density (OD). (d) Serologic phenotype of antibody presence in seropositive participants (e) US Map showing seropositivity in six regions surveyed: Northeast = ME, NH, VT, MA, NY, CT, RI, PA, NJ, 7.5% (95% CI: 3.7 – 11.3%); Midwest = MN, IA, WI, IL, IN, MI, OH, 1.6% (95% CI: 0.06-2.3%); Mid-Atlantic = MD, DE, DC, VA, WV, KY, TN, NC, SC, GA, 8.6% (1.3 – 15.8%); South/Central = FL, MS, AL, LA, AR, MO, KS, OK, 3.0% (1.2 – 4.5%); Mountain/Southwest = TX, NM, AZ, CO, UT, WY, NE, SD, ND, MT, ID, 4.5% (0.09 – 7.9%); West/Pacific = WA, OR, NV, CA, AK, HI, 1.9% (0.02 – 3.2%). One person in diagram represents 100 participants, orange represents weighted prevalence estimate within the geographic region.

### Statistical Analysis

The previously described iterative quota sampling continuously matched the proportion of people in the study with the census estimated proportion of people in the country on 6 variables (**Table 1, Figure 1**). This ensured that each periodic sample of participants over the course of the study were representative, and the time effects of the pandemic were approximately independent of those 6 variables. Each participant was asked demographic and health-related questions that matched ones on the Behavioral Risk Factor Surveillance System (BRFSS) survey, a large probability-based national survey^19^. Responses to those matching questions were used with BRFSS survey data to adjust estimators to account for important criteria that may be related to both selection probability and seropositivity but were not accounted for in the quota sampling. Those adjusted estimators used weighting based on the propensity of being a quota sample versus a BRFSS sample participant and poststratification to US census data. It additionally accounted for sensitivity and specificity. Confidence intervals were calculated for the final seroprevalence estimates accounting for both the variability of the weighting and of the sensitivity and specificity adjustment. The ratio of undiagnosed cases over diagnosed cases was estimated as the final seroprevalence estimate times a factor calculated from the daily national population and diagnosed cases. For more methods and details see Section 3 of the **Supplementary Materials**.

## RESULTS

### Enrollment and Demographic Representation

Recruitment took place from April 1, 2020 until August 4, 2020. During that time 11,283 participants were enrolled from a pool of 241,424 volunteers. Of these participants, 214 had blood collected via venipuncture and 11,069 were sent microsamplers. Over 80% of the microsamplers were returned (9,089 participants). Ultimately 9,028 participant samples were analyzed via ELISA for presence of SARS-CoV-2 antibodies. Of those, 8,058 participants had complete clinical questionnaire data and were included in the weighted analysis (**Figure 1**). The majority (>88%) of sample collection occurred within the 11-week period between May 10^th^ to July 31^st^, 2020 (**Supplemental Fig. 3**). The six major demographic factors used in participant selection are summarized in **Table 1**. Participant sampling was highly representative of the U.S. population. When expanded to include the additional 10 demographic or health related factors captured by the BRFSS, many factors were well matched, but there were some differences: our sample population was more highly educated, employed, and had better access to healthcare (**Table 1**).

**Figure 3:**
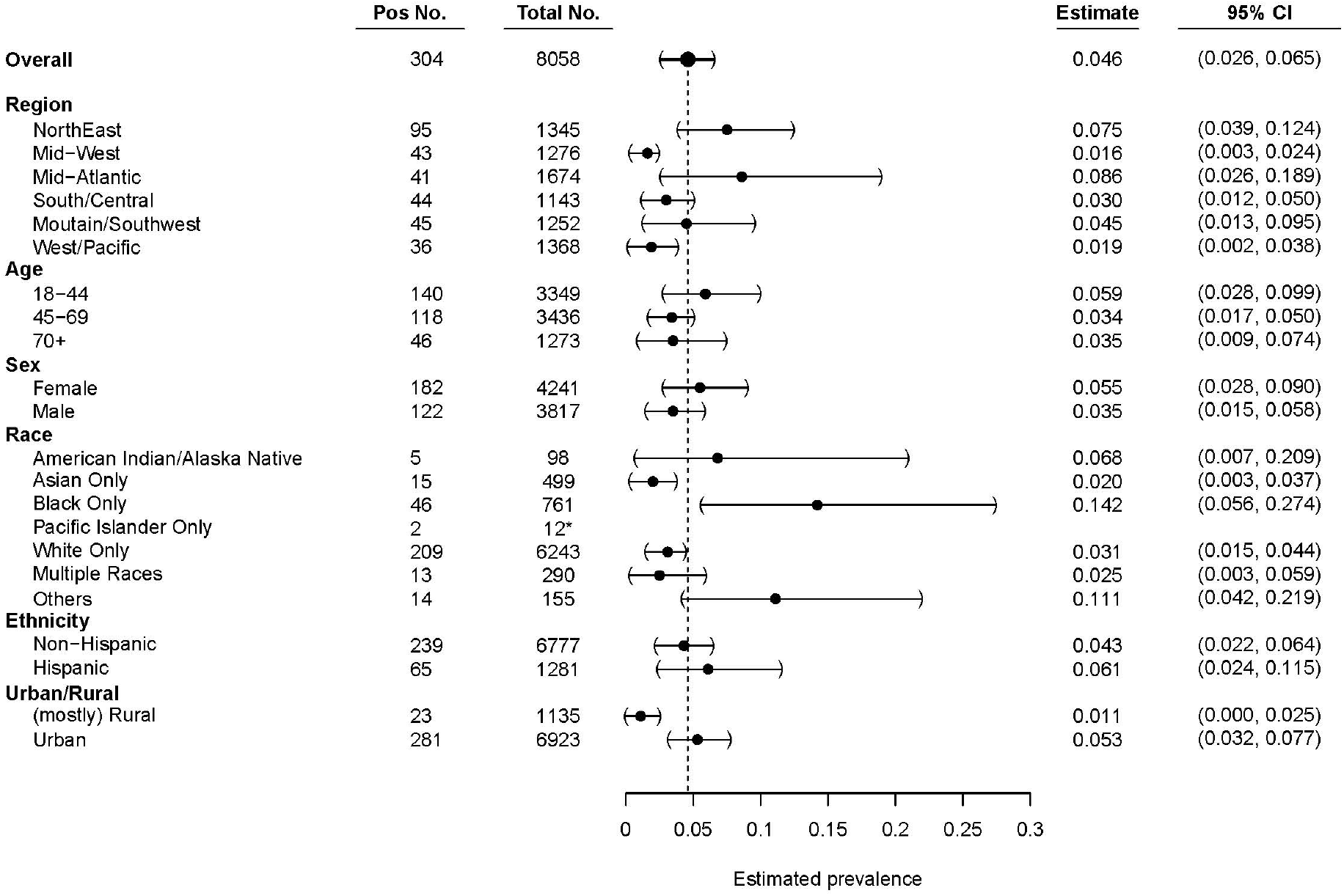
Undiagnosed seroprevalence in main demographic categories. Six main categories utilized during quota-based sampling: region, age, sex, race, ethnicity, and urban/rural. Seropositivity estimates of samples that had a full clinical questionnaire completed and successful sampling. Data are weighted estimates ± 95% confidence intervals. Dashed line = weighted national seroprevalence estimate. ^*^ = n value too low to make proper weighted estimate, raw positivity displayed.

### Estimates of Seroprevalence

There were 304 seropositive participants in the analysis set (**Figure 2a,b**). This gave a weighted estimate of 4.6% of the undiagnosed adults in the U.S. population that were seropositive for SARS-CoV-2 (95% CI: 2.6% to 6.5%, *n* = 8058 complete testing and survey). Using this average rate over the study period, we estimate that there were 4.8 undiagnosed cases per each diagnosed case over the course of the study (95% CI: 2.8, 6.8). In seropositive participants, 36.51% were IgG^+^IgM^+^IgA^+^, 28.29 % were IgG^+^IgM^-^IgA^+^, 17.11% were IgG^+^IgM^-^IgA^-^, 13.16 % were IgG^+^IgM^+^IgA^-^, 4.28 % were IgG^-^IgM^+^IgA^-^, and 0.66 % were IgG^-^IgM^+^IgA^+^ (**Figure 2a-c, Supplemental Fig. 4**).

**Figure 4:**
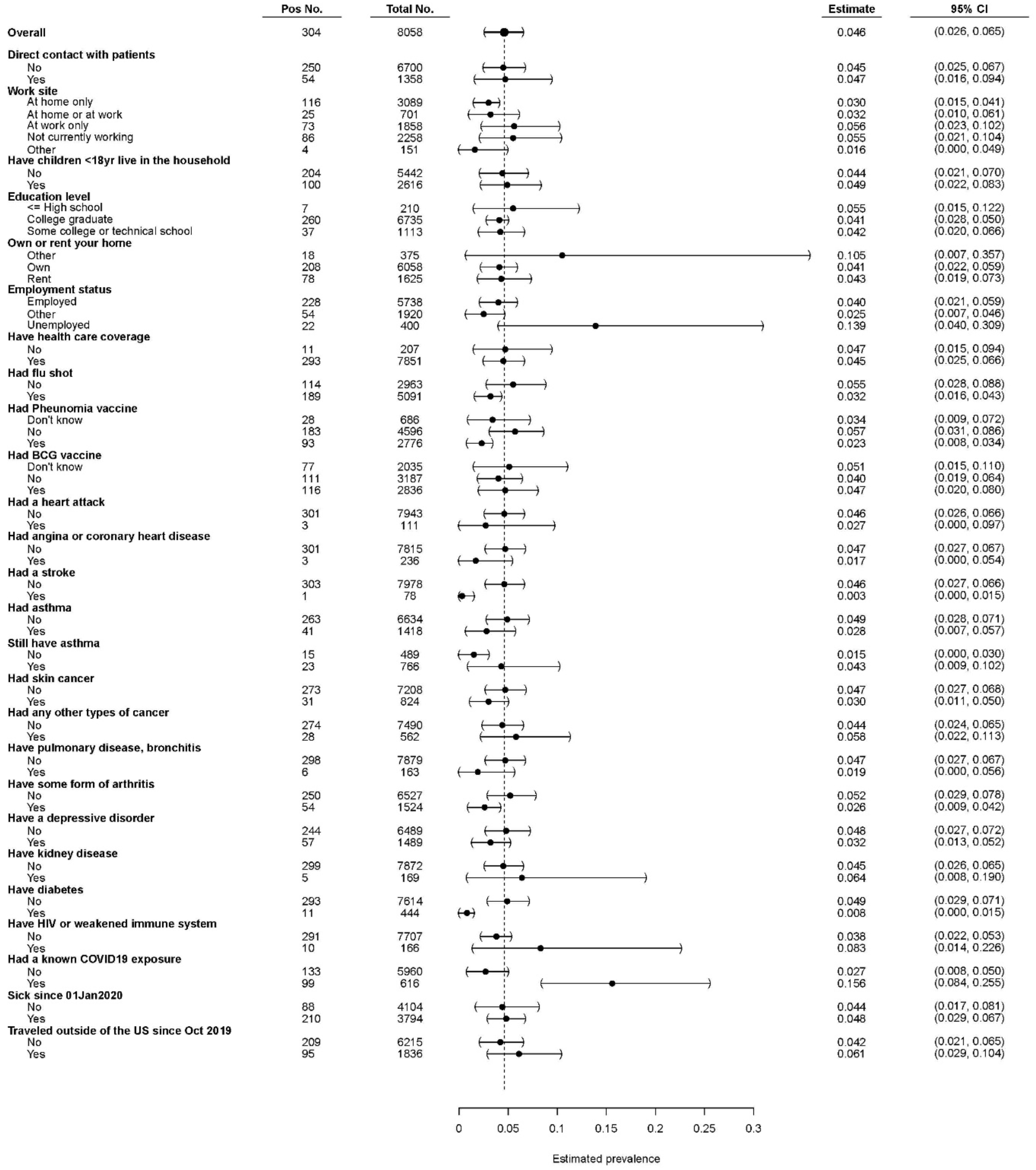
Seroprevalence estimates of health and behavioral traits. Seropositivity estimates of samples that had a full clinical questionnaire completed and successful sampling. Data are weighted estimates ± 95% confidence intervals. Dashed line = weighted national seroprevalence estimate.

We found regional variations of seroprevalence estimates across the US (**Fig. 2d, 3**). The Northeast and Mid-Atlantic Regions showed the highest rates of seropositivity whereas the lowest in the Midwest. Urban areas were estimated to have higher levels of seropositivity (5.3%) compared to rural areas (1.1% seropositivity) at the time samples were collected. Estimates of seroprevalence were calculated for other demographic subgroups (**Figure 3**). The youngest age group, 18-44, had the highest estimated seropositivity (5.9%). Estimated seroprevalence for females was 5.5% and 3.5% in males. The seroprevalence estimate for Black/African Americans was highest at 14.2% followed by participants who self-identified as other/unlisted race (11.1%), American Indian/Alaska Native (6.8%), followed by White/Caucasian (2.5%), while those identifying as Asian displayed the lowest seroprevalence estimate (2.0%).

Participants who reported a known exposure to a SARS-CoV-2-infected individual had a higher seroprevalence estimate (15.6%) compared to those who did not (2.7%). In comparison to the national average (4.6%), those that worked from home had a lower seropositivity estimate of 3.0%. Those who reported prior vaccination (influenza 3.2% and/or pneumonia 2.3%) had a lower likelihood for undiagnosed seropositivity. Those who had health conditions associated with poor outcomes in SARS-CoV-2 infection, including coronary heart disease, asthma, and diabetes, displayed lower rates of seropositivity (**Figure 4**). Other health conditions were also correlated with a decreased seropositivity rate such as skin cancer, stroke, or arthritis.

## DISCUSSION

This study demonstrates that spread of the SARS-CoV-2 virus in the US during the first six months of the pandemic was more widespread than has been suggested by data reporting diagnostic test-confirmed cases. Similar to responses to other respiratory viruses, such as influenza, many individuals develop asymptomatic or mild disease that is not medically attended and therefore never diagnosed. Our findings indicate that there are nearly five individuals with a previous asymptomatic infection for every diagnosed case. Furthermore, patterns of our seroprevalence data match well with those of diagnosed cases reported during a similar timeframe.^20^ For example, the greater seropositivity estimated in densely populated urban areas follows the observed initial spread of SARS-CoV-2. In comparison to the national average, we found that the Midwest, South, and West had lower seroprevalences during the study timeframe, which preceded a substantial increase in infections in these regions detected by viral testing.

Our data suggest that the youngest age group had the highest undiagnosed seroprevalence, which is consistent with observations that they display less severe symptoms than older patients^21^. We also found higher undiagnosed seroprevalence in females, possibly suggesting a higher risk for asymptomatic disease. Participants with chronic diseases that are more likely to be associated with severe clinical manifestations of COVID-19, including diabetes, heart disease, and asthma, had a lower prevalence of asymptomatic SARS-CoV-2 infection in comparison to the national average. Those with known exposure to SARS-CoV-2 infected individuals had a higher estimated incidence of undiagnosed seropositivity. We also found that Black, African American, and Hispanic participants had higher undiagnosed seropositivity, correlating with national data on disease burden in these sub-groups.

This study is the first to report a representative sample across the US and to evaluate regional, demographic and socioeconomic differences in the prevalence of asymptomatic SARS-CoV-2 infection. In contrast, other reports of seroprevalence data focus on a specific group of individuals or geographic location^22^. Our results provide new insight into the spread of SARS-CoV-2. Our estimate of the national undiagnosed exposure rate provides information on the scope of infection during the first six months of the pandemic. This work extends findings from smaller foundational studies of limited populations^23-37^ by generating an accurate estimate of nationwide and subgroup prevalence.

Our results estimate that there are approximately 4.8 undiagnosed cases (95% CI 2.76-6.81) for every identified case of COVID-19, suggesting a potential 16.8 million undiagnosed cases by mid-July 2020 in addition to the reported 3 million diagnosed cases in the United States. These data suggest a higher level of infection-induced immunity exists in the population and the size of those with this immunity is even greater now as the virus continued to spread in the months since this study was performed. Further long-term studies of immunity in the population will be necessary to further understand durability of response to the vaccine versus infection, how infection-induced immunity impacts vaccine response and performance, and if herd immunity can play a role in controlling SARS-CoV-2 spread. In addition, further subgroup analysis of our data will be useful in clarifying the spread of disease in the presence of public health measures and how we may be able to refine and further target those measures in the future.

### Limitations

Although we were able to recruit a cohort with demographics representative of the general US population, our study has several limitations. First, although extensive statistical adjustments were made, our study cohort is based on a non-random volunteer sample which can have selection bias. However, many traditional random sampling studies using probability sampling design have very low response rates, calling into question the advantages of that practice^38,39^. Our study population also exhibited some differences from the general US population, such as higher education level and access to healthcare that had to be adjusted for with statistical weighting. We utilized both census and behavioral data to weight our results though it is possible that there are variables associated with disease transmission that are not accounted for in our weighting.

## CONCLUSIONS

These data suggest a much larger spread of the COVD-19 pandemic than originally thought and have implications in basic understanding of SARS-CoV-2 spread, epidemiologic characteristics of its spread and prevalence in different communities, and potential impact on decisions involved in vaccine rollout. Continued large-scale surveillance of SARS-CoV-2 immunity is in progress, discriminating infection-based and vaccine-induced antibody responses, and mathematical models will be generated to understand the pandemic, vaccine performance, public health measure efficacy, and providing insight for our approach to handling the next virus with pandemic potential.

## Data Availability

Data are available upon reasonable request to corresponding authors on a case-by-case basis as the clinical study is still ongoing.

## ACKNOWLEDGEMENTS

Firstly, we would like to thank the participants in this study, without whom it would not have been possible. The authors would like to acknowledge Dr. Corbett and Dr. Graham of the NIAID VRC for their generous donation of coronavirus spike expression plasmid, and Dr. Aaron Schmidt, J Feldman, BM Hauser and T M Caradonna of the Ragon Institute of MGH, MIT, and Harvard for their donation of their RBD expression plasmid. We thank John Crissey and the CRIMSON team (NIAID, NIH) for their assistance and contributions to the study and data management. We thank Dr. Jason McLellan for scholarly discussions regarding the spike S-2P construct. We thank the CCTS Clinical Research Support Program (A. Delbridge, L. Dukes), UAB School of Public Health and its Survey Research Unit (L. Battle, J. Carson, M. DeRamus, T. Fields, T. Graham, T. Jackson, E. Pruitt, A. Underwood, P. Wolff), the University of Pittsburgh Clinical and Translational Science Institute staff and leadership (J. Avolio, L. Bash, S. Clayton, M. Cristinziano, K. Daw, C. Fascetti, E. Gyurisin, J. Huwe, N. Jones, D. Mathias, S. Mathias, A. Mykita, B. Petersen, M. Phillips, C. Rush, E. Shepherd, S. Shetty, A. Socci, L. Stearns, S. Ugbomah, K. Underwood, L. Yasko) and University of Pittsburgh Information Technology (D. McGaughey, S. Ritzman, T. Smith) for their contributions to this work. We would like to thank Mr. Denzel Bernard for his assistance with sample delivery, and Dr. Stephanie Ford-Scheimer for organizational assistance. This research was supported in part by the Intramural Research Program of the NIH, including the National Institute for Biomedical Imaging and Bioengineering, the National Institute of Allergy and Infectious Disease, and the National Center for Advancing Translational Sciences. This project has been funded in part with Federal funds from the National Cancer Institute, National Institutes of Health, under contract number HHSN261200800001E, 75N91019D00024, Task Order No. 75N91019F00130, Clinical and Translational Science Awards Program grants UL1TR003096 (UAB) and UL1TR001857 (University of Pittsburgh). The content of this publication does not necessarily reflect the views or policies of the Department of Health and Human Services, nor does mention of trade names, commercial products, or organizations imply endorsement by the U.S. Government. <u>Disclaimer</u>: The NIH, its officers, and employees do not recommend or endorse any company, product, or service.

