## Supplementary Materials for "Mapping a Pandemic: SARS-CoV-2 Seropositivity in the United States"

#### SUPPLEMENTARY FIGURES AND LEGENDS

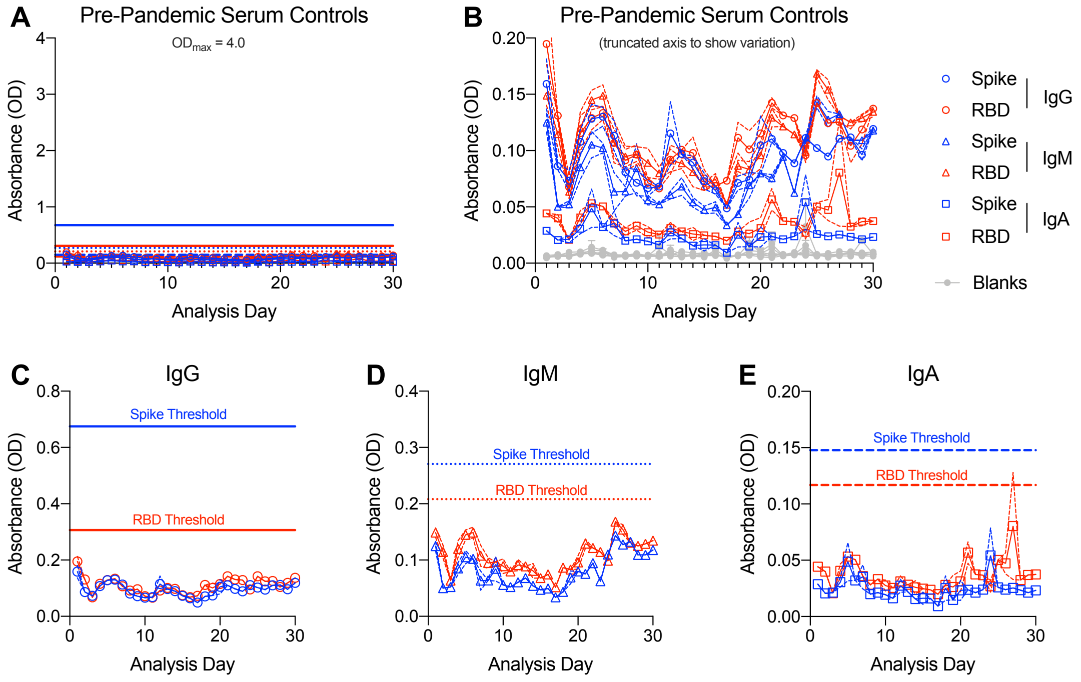


**Supplemental Figure 1: Pre-pandemic sera controls in 30 days of sample analysis**. (a) Pre-pandemic sera controls for spike (blue) and RBD (red) ELISAs. (b) Truncated axis to show variation not visible with full axis. (c) IgG controls with truncated axis to show variation. (d) IgM controls with truncated axis to show variation. (e) IgA controls with truncated axis to show variation. Spike = blue, RBD = red, Blank = grey. IgG = open circles, IgM = open triangles, IgA = open squares. Data are means ± SD of plates run per day (thin dashed lines around data points), *n* = 10 per day.

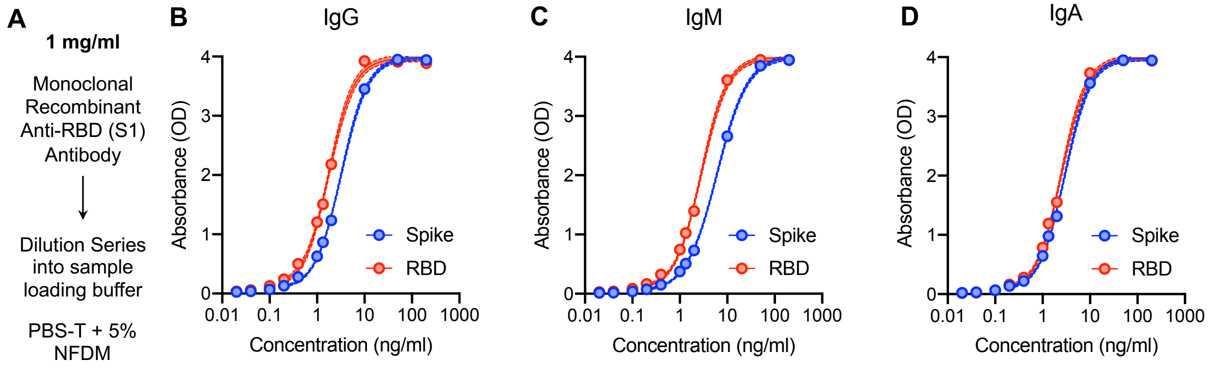


E

| | IgG OD | IgG ng/ml | IgM OD | IgM ng/ml | IgA OD | IgA ng/ml |
|---|---|---|---|---|---|---|
| **2SD Threshold** | | | | | | |
| Spike | 0.67472084 | 2.13558269 | 0.2081296 | 2.85143534 | 0.09136142 | 0.95060312 |
| RBD | 0.30626822 | 0.88452737 | 0.27055366 | 1.37172674 | 0.11278165 | 0.89259726 |
| **3SD Threshold** | | | | | | |
| Spike | 0.90991654 | 2.35835233 | 0.27006523 | 3.07883151 | 0.11674446 | 1.07096164 |
| RBD | 0.38678635 | 0.95962154 | 0.35802882 | 1.50820606 | 0.1475528 | 0.99448861 |

**Supplemental Figure 2: Recombinant antibody dilution curves.** (a) Dilution curve setup. Recombinant antibody dilution curve plotted against resulting absorbance (optical density, A450 – A650) on ELISA for (b) IgG, (c) IgM, (d) IgA. (e) Threshold values translated into quantitative concentration value for recombinant antibody at two and three standard deviations above the mean of the archival negative controls. (See Klumpp-Thomas et al, *Nature Comms* 2020)

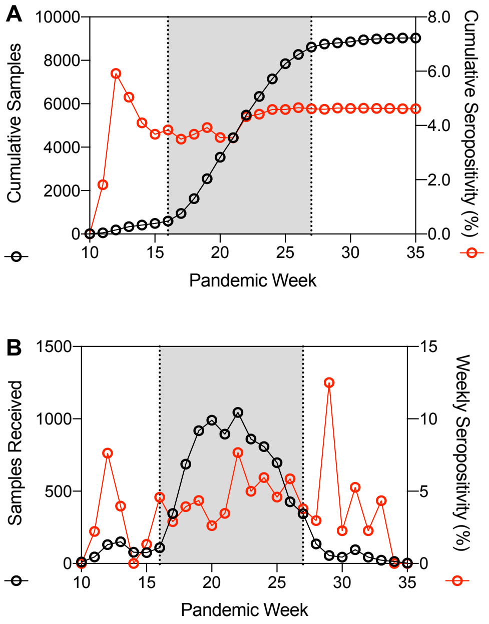


**Supplemental Figure 3: Collection timeframe plotted against seropositivity.** (a) Cumulative sample collection (black) and cumulative unweighted seropositivity (red) over the course of the study. (b) Weekly samples received (black) and weekly unweighted seropositivity (red) over the course of the study. Pandemic week: week of the US pandemic starting with week 1 = the week of the first official reported case in the United States (January 22nd). Grey shaded area = window in which the majority of samples were collected between May 31st and July 14th, 2020. A majority of the convenience sample recruitment occurred between weeks 10 and 13 of the pandemic.

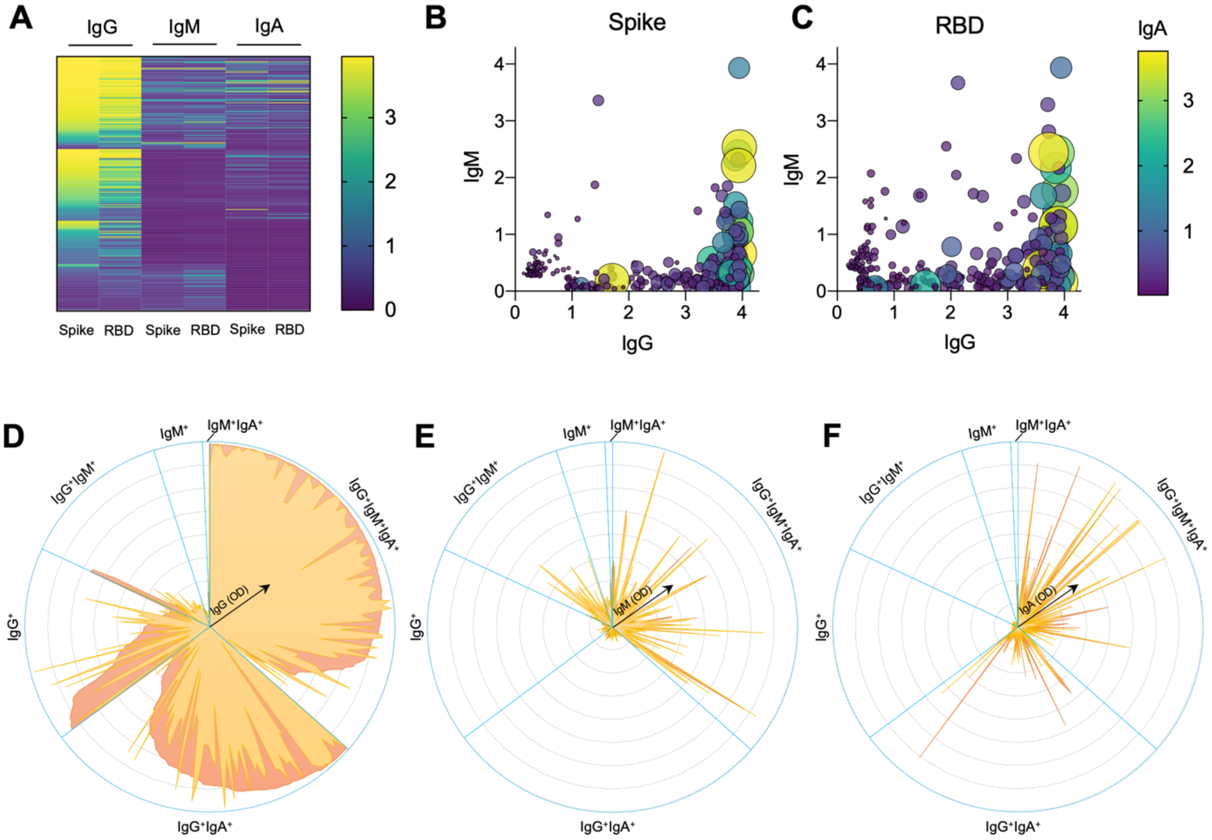


**Supplemental Figure 4: Raw ELISA data and antibody profiles of seropositive participants**. (a) Heatmap showing optical density (OD) values for the 304 seropositive participants. (b) Anti-spike antibodies with OD displayed for IgG (x-axis), IgM (y-axis) and IgA (point size/color). (c) Anti-spike antibodies with OD displayed for IgG (x-axis), IgM (y-axis) and IgA (point size/color). (d-f) OD for spike (orange) and RBD (yellow) for each sample, separated by seropositivity determination for (d) IgG, (e) IgM and (f) IgA.

### SUPPLEMENTAL TABLES AND LEGENDS

**Supplementary Table 2: Sigmoidal 4 Parameter Logistic Modeling of Recombinant Antibody Dilution Curves in Sample Loading Buffer**

| IgA | | |
|---|---|---|
| **Best-fit values** | | |
| Bottom | 0.04889 | 0.04667 |
| Top | 3.985 | 3.993 |
| IC50 | 2.954 | 2.475 |
| HillSlope | 1.623 | 1.639 |
| logIC50 | 0.4704 | 0.3936 |
| Span | 3.936 | 3.946 |
| **95% CI (asymptotic)** | | |
| Bottom | 0.02475 to 0.07303 | 0.01502 to 0.07833 |
| Top | 3.944 to 4.025 | 3.943 to 4.043 |
| IC50 | 2.836 to 3.072 | 2.359 to 2.591 |
| HillSlope | 1.544 to 1.702 | 1.528 to 1.750 |
| logIC50 | 0.4527 to 0.4875 | 0.3728 to 0.4134 |
| Span | 3.886 to 3.986 | 3.883 to 4.010 |
| **Goodness of Fit** | | |
| Degrees of Freedom | 44 | 44 |
| R squared | 0.9989 | 0.9982 |
| Adjusted R squared | 0.9988 | 0.9981 |
| Sum of Squares | 0.1265 | 0.2076 |
| **Replicates test for lack of fit** | | |
| SD replicates | 0.02632 | 0.02416 |
| SD lack of fit | 0.1127 | 0.1527 |
| Discrepancy (F) | 18.34 | 39.96 |
| P value | <0.0001 | <0.0001 |

| IgM | | |
|---|---|---|
| **Best-fit values** | | |
| Bottom | 0.02553 | 0.06548 |
| Top | 4.02 | 3.996 |
| IC50 | 6.072 | 2.834 |
| HillSlope | 1.342 | 1.574 |
| logIC50 | 0.7833 | 0.4524 |
| Span | 3.995 | 3.93 |
| **95% CI (asymptotic)** | | |
| Bottom | 0.004449 to 0.04660 | 0.03690 to 0.09406 |
| Top | 3.977 to 4.063 | 3.948 to 4.044 |
| IC50 | 5.827 to 6.317 | 2.703 to 2.966 |
| HillSlope | 1.291 to 1.392 | 1.483 to 1.664 |
| logIC50 | 0.7654 to 0.8005 | 0.4318 to 0.4721 |
| Span | 3.944 to 4.045 | 3.872 to 3.989 |
| **Goodness of Fit** | | |
| Degrees of Freedom | 44 | 44 |
| R squared | 0.9991 | 0.9985 |
| Adjusted R squared | 0.999 | 0.9984 |
| Sum of Squares | 0.09551 | 0.1721 |
| **Replicates test for lack of fit** | | |
| SD replicates | 0.04303 | 0.02226 |
| SD lack of fit | 0.06006 | 0.1388 |
| Discrepancy (F) | 1.949 | 38.91 |
| P value | 0.0824 | <0.0001 |

| IgG | | |
|---|---|---|
| **Best-fit values** | | |
| Bottom | 0.05631 | 0.07779 |
| Top | 3.99 | 3.977 |
| IC50 | 3.253 | 1.778 |
| HillSlope | 1.559 | 1.628 |
| logIC50 | 0.5123 | 0.25 |
| Span | 3.933 | 3.899 |
| **95% CI (asymptotic)** | | |
| Bottom | 0.03375 to 0.07887 | 0.03468 to 0.1209 |
| Top | 3.951 to 4.029 | 3.917 to 4.037 |
| IC50 | 3.127 to 3.380 | 1.695 to 1.862 |
| HillSlope | 1.492 to 1.626 | 1.481 to 1.775 |
| logIC50 | 0.4951 to 0.5289 | 0.2291 to 0.2700 |
| Span | 3.886 to 3.981 | 3.820 to 3.978 |
| **Goodness of Fit** | | |
| Degrees of Freedom | 44 | 44 |
| R squared | 0.999 | 0.9971 |
| Adjusted R squared | 0.9989 | 0.9969 |
| Sum of Squares | 0.1103 | 0.3397 |
| **Replicates test for lack of fit** | | |
| SD replicates | 0.02497 | 0.04346 |
| SD lack of fit | 0.1048 | 0.1843 |
| Discrepancy (F) | 17.61 | 17.99 |
| P value | <0.0001 | <0.0001 |

x = concentration of recombinant antibody in sample incubation buffer

**Supplemental Table 1: Sigmoidal 4 Parameter Logistic Modeling of Recombinant Antibody Dilution Curves**

#### Statistical Supplement & Methods

1 Statistical Methods

##### 1.1 Overview

The primary purpose of this study is to estimate the prevalence of seropositivity to the SARS-CoV-2 virus antibody among the target population of adults 18+ years old living in the US who had never been diagnosed during the early phases of the pandemic (enrollment started in late March and ended in late September, 2020). Because of the changing dynamics of the pandemic, there was an urgency to begin and complete a study rapidly; therefore, the study relied on using volunteer samples rather than using a random sample. Initially, a convenience sample of 593 participants from the Washington, DC metropolitan area were enrolled. Later the scope of the study expanded, and the samples were collected through a quota sampling process from the volunteer registry. Specifically, a very large national pool of volunteers (241,424, see Figure 1) filled out a brief screening questionnaire with demographic data, then a quota sample of individuals were selected from that pool so that their distribution on 6 variables matched the distribution of these same variables in the US census. In Section 3.2 we describe the US census data used, and in Section 3.4 we describe the quota sampling algorithm. Although the quota sampling was a random sample with known probabilities of selection from the pool, the pool of volunteers itself has potentially large selection bias. Selection biases that are not accounted for by the quota sampling variables can also occur (see Figure 1 of the main paper). To adjust for these selection biases, we use the Behavioral Risk Factor Surveillance System (BRFSS), which is a US representative survey (Section 3.3), to create a weight for each observation in the analytic data set. The weight is created so that the weighted distributions of selected variables in our study agrees with the distributions of the BRFSS (Section 3.5). After obtaining the adjusted estimates with associated standard errors, we further correct the prevalence estimates for the sensitivity and specificity of the antibody assays (including the variability of the sensitivity and specificity estimates) to get confidence intervals on the adjusted prevalence estimator (Section 3.6). Finally, we describe other statistics calculated in the paper in Section 3.7.

##### 1.2 US Census Data

To determine the proportions of volunteers selected in the quota sample (by the 6 variables), we used US Census data for 2018 (US Census Bureau, 2020). This data was the most current census data available at the planning stage in April 2020 that had race and ethnicity information by county. We additionally used age group and sex by county information from the 2018 estimates. The catchment area was the 50 states plus Washington DC. We divided the catchment area into sets of states or DC, defining 6 roughly equal sized (by total population) regions, to facilitate division of administrative workload for the data collection. The total populations for the regions were between about 50 million to 56 million. The 6 regions are listed in Statistical Table 1.

Statistical Table 1: Definition of the Geographic Regions

| Region Number | Total (including children) 2018 Population Size (in millions) | State or District |
|---|---|---|
| 1 | 56.1 | Connecticut, Maine, Massachusetts, New Hampshire, Rhode Island, Vermont, New Jersey, New York, Pennsylvania |
| 2 | 55.7 | Illinois, Indiana, Michigan, Ohio, Wisconsin, Iowa, Minnesota |
| 3 | 55.3 | Delaware, District of Columbia, Georgia, Maryland, North Carolina, South Carolina, Virginia, West Virginia, Kentucky, Tennessee |
| 4 | 49.8 | Kansas, Missouri, Florida, Alabama, Mississippi, Arkansas, Louisiana, Oklahoma |
| 5 | 53.8 | Nebraska, North Dakota, South Dakota, Texas, Arizona, Colorado, Idaho, Montana, New Mexico, Utah, Wyoming |
| 6 | 56.5 | Nevada, Alaska, California, Hawaii, Oregon, Washington |

The quota sampling was designed to match on the marginal distributions of 6 variables: state/district, age group, sex, ethnicity, race, and urban/rural (see Statistical Table 2). The only quota variable that needs further explanation is age. The census data is listed by age in 5-year age groups. To obtain the adult population totals we used the populations from the age groups for ages 20-24 and older, and also included 2/5 of the 15-19 age group to represent ages 18 and 19. The other variable levels are as defined by the US Census (US Census, 2020). For our analyses that combine the census data with the BRFSS data and the seroprevalence data, the urban/rural was slightly modified to harmonize the data sets (see next section), and additionally in the BRFSS data, race had a self-defined "other" category.

Statistical Table 2: Quota variables

| Quota Variable | Definition of categories |
|---|---|
| **Age (3 categories)** | 1 = 18-44 years old<br>2 = 45-69 yrs<br>3 = 70+ yrs |
| **Sex (2 categories)** | Male, female |
| **Ethnicity (2 categories)** | Hispanic, non-hispanic |
| **Race (6 categories)** | WA=White alone<br>BA=Black or African American alone<br>IA=American Indian and Alaska Native alone<br>AA= Asian alone<br>NA=Native Hawaiian and Other Pacific Islander alone<br>TOM= Two or More Races |
| **Urban/Rural (3 categories)** | mostly urban<br>mostly rural<br>completely rural |
| **State/district (51 categories)** | 50 states + Washington DC |

1.3 Behavioral Risk Factor Surveillance System (BRFSS) Data

At the time of the study design the most recently available BRFSS data was the 2018 data (CDC, 2018). We chose questions from the BRFSS to include in our seroprevalence study that are possibly related to the selection probability for volunteering into our study (see study protocol for questions). For the adjustment we used the first 4 out of the 6 quota sampling variables in Statistical Table 2, plus modifications of the other 2 quota variables. The urban/rural variable for the BRFSS is slightly different than the US Census designations and follows the urban counties vs. rural counties based on 2013 Office of Management and Budget standards for defining metropolitan statistical areas (Ingram and Franco, 2013). For the sixth quota variable, we collapsed the state/district quota variable into regions (see Statistical Table 1). Additionally, 10 more variables were created from either a single question or combination of questions in both BRFSS and our seroprevalence survey (Statistical Table 3) for adjustment. Questions were combined if the questions were on a similar topic.

Statistical Table 3: Variables used for selection bias adjustment

| **Question Class** | **Topic** | **Question code for 2018 BRFSS** |
|---|---|---|
| Quota questions | sex | C08.01 |
| | age | C08.02 |
| | ethnicity | C08.03 |
| | race | C08.04 |
| | Urban/rural (slightly different than census) | (see Ingram and Franco, 2013) |
| | region (instead of state/district) | (use Statistical Table 1 of this document) |
| BRFSS Adjustment Questions | # children in household | C08.16 |
| | Education | C08.07 |
| | homeowner status | C08.08 |
| | employment | C08.15 |
| | Health care coverage | C03.01 |
| | any cardio-vascular disease? | C06.01, C06.02, C06.03 |
| | any asthma or lung problems? | C06.5, C06.08 |
| | any cancer (besides skin cancer) or immunological problems? | C06.07, C06.09 |
| | any diabetes? | C06.12 |
| | Flu shot in last 12 months or ever had pneumococcal vaccine? | C11.01, C11.04 |

The BRFSS is a very large probability-based random sample of the United States plus its territories. The BRFSS was designed so that BRFSS observations weighted by sampling weights give representative estimates for the entire US. We checked how well the BRFSS weighted estimates are for our quota sample variables compared to the census values for the quota variables. We see in Statistical Table 4 that they generally match well except for urban/ rural and raceGrp3 where there are some discrepancies.

Statistical Table 4: Comparison of weighed proportions from the BRFSS to the proportions from the US Census

| | 2018 BRFSS | | | 2018 Census | |
|---|---|---|---|---|---|
| **Region** | **n** | **%** | **Weighted (%)** | **N** | **%** |
| 1 | 91307 | 21.19 | 17.6 | 44,427,073 | 17.52 |
| 2 | 67110 | 15.57 | 16.97 | 43,187,427 | 17.03 |
| 3 | 80979 | 18.79 | 16.91 | 42,888,136 | 16.91 |
| 4 | 60482 | 14.03 | 15.35 | 38,959,350 | 15.36 |
| 5 | 86204 | 20 | 15.89 | 40,394,958 | 15.93 |
| 6 | 44866 | 10.41 | 17.27 | 43,766,713 | 17.26 |
| **Agegr3** | | | | | |
| [18,44] | 125081 | 28.59 | 46.00 | 117,288,129 | 46.24 |
| [45,69] | 207749 | 47.49 | 39.84 | 100,991,228 | 39.82 |
| [70+] | 104605 | 23.91 | 14.17 | 35,344,300 | 13.94 |
| **Sex** | | | | | |
| Male | 197411 | 45.24 | 48.66 | 123,556,702 | 48.72 |
| Female | 238911 | 54.76 | 51.34 | 130,066,955 | 51.28 |
| **NCHS.Urban.Rural*** | | | | | |
| Urban county | 365714 | 84.9 | 93.48 | 220,499,070 | 86.94 |
| Rural county | 65234 | 15.1 | 6.52 | 33,124,588 | 13.06 |
| **RaceGrp3** | | | | | |
| White only | 345710 | 81.00 | 73.41 | 197,029,928 | 77.69 |
| Black only | 37862 | 8.87 | 12.90 | 32,690,602 | 12.89 |
| Others | 43219 | 10.13 | 13.69 | 23,903,128 | 9.42 |
| **Hispanic** | | | | | |
| Yes | 36941 | 8.53 | 17.06 | 41,179,568 | 16.24 |
| No | 395931 | 91.47 | 82.94 | 212,444,090 | 83.76 |

*The urban/rural designation for NCHS (used with BRFSS data) was defined differently than in the census, and the list of US counties and county-equivalent entities and their urbanization levels (Table V in https://www.cdc.gov/nchs/data/series/sr_02/sr02_166.pdf) is used for Census counts.

#### 1.4 Quota sampling algorithm

The initial convenience sample of 593 (and their characteristics) was used to start the algorithm for the quota sampling, to draw participants from the large volunteer pool of 241,424 (see Figure 1). This section describes that algorithm.

There are three data sets used in this algorithm.

1. The target data is created from the census data (Section 1.2). First we calculate the proportion of people in the census within each cross-classification of Sex, Age group, Ethnicity, Race, State/District, and Rural/Urban status (based on county); which gives 9936 unique cells (because for example, not all

states/district have all three of the Rural/Urban categories). Then we multiply the proportion for each cell by 10,000, the original planned study size. Many of these cells will have values less than 1.

2. The enrolled data is collected from the set of individuals who have been consented and completed their demographic survey.
3. The volunteer pool data is the set of individuals who have volunteered and filled out a screening survey with demographic information.

Over the course of the study some individuals in the volunteer pool were removed as they are invited to participate and potentially move to the enrolled data. The algorithm is applied iteratively, and it decides who to invite based on the current volunteer pool and the current enrolled sample, using the target data.

In the first iteration of the algorithm, the convenience sample sets the sampling probabilities to select participants to invite.

For the subsequent periodic invitations, the quota algorithm does the following:

1. Update the target data by subtracting out the number already enrolled within each of the 9936 cells (i.e., the sub-classifications by each level of the combination of 6 variables that are present in the census). At this step, we keep negative values for cell targets, because most of the sub-classifications have targets < 1, so even getting one person into the group will make the target go negative; however, we are not focused on the specific cells and instead look at the marginal proportions for each of the 6 quota variables (Statistical Table 2). As an example of the marginal proportion matching, we want to match the target proportion of males and females, but the individual cells do not necessarily have to (nor even can) match the male and female proportions within each county and age group and race.
2. Eliminate anyone from the volunteer pool who has previously been invited to participate. Calculate the number remaining in the volunteer pool for each of the 9936 cells.
3. For each of our 9936 cells, then, we can designate a probability sample as the updated target number within that cell divided by the pool of volunteers available in the corresponding cell. If the probability sample is negative, then we set the probability sample to zero.
4. We also calculated the shortfall for each of the 9936 cells by taking the difference between the updated target sample needed and the available volunteer pool, keeping all positive values and setting all other values to 0. The shortfall was then summed across each of the 6 quota variables and divided by the corresponding updated targets, to determine the shortfall in each category are needed. For example, for the state/district variable, sum within the 51 categories to see how many people are still needed from each state/district. We analogously take the marginal sum of shortfalls across each of the 6 quota variables, to find out how many are still available in the volunteer pool. For large values of the shortfall, we increase the sampling probability for that category by 10-200% depending on how large the shortfall is. For example, if we are deficient in proportion of people aged 70+ in the volunteer pool, we can increase the sampling probability by a constant factor for all cells representing that age group.
5. The regions are divided into 3 administrative groups: two are the Clinical and Translational Science Awardees from University of Alabama at Birmingham and the University of Pittsburgh (regions 1, 2, 4, 5, and 6), and the third group is NIAID (region 3). Based on the availability of personnel, we estimate the number of people to be called each day for each administrative group. We then take sample of size $s_j$ from our volunteer pool, where s1,s2, and s3 are proportional to the sizes needed for the 3 administrative groups. Using the previously calculated sampling probabilities (Step 4 above) we sample a data set for each of the 3 administrative groups and combine them to get a simulated sampling for the full study. We repeat this sampling 20,000 times so that we have 20,000 sets of samples of size s=s1+s2+s3.
6. To select the "best" sample (out of the 20,000 available), we calculate the marginal proportions of our sample in each group and compare it to the desired breakdown according to our updated targets. For example, we will calculate the percent male/female in each of the 20,000 samples and compare that to the percent male/female according to our updated targets. For each quota variable, we calculate a weighted sum of the square of the differences between the simulated proportion (for one simulated data set) and the target proportion. Let $k_j$ be the number of categories for the $j^{th}$ quota variable. Then the weighted sum of squared

differences sums the first $k_j$-1 of the differences and divides by ($k_j$-1) so that each variable is given equal importance. For example, we sum the first 50 squared deviations of the 51 states/district and divide by 1/50 and for sex we just calculate the squared deviations of the males (and divide by 1). We select the simulated sample with the lowest weighed sum of squared differences and use that for our invitation list.

#### 1.5 BRFSS adjustment for selection bias

Although probability sampling has been the "touchstone for good survey practice", recent survey samples have had very low response rates calling into question that touchstone practice (Elliott and Valliant, 2017, Hill, 2020). Further, due to the need to quickly field the survey in the early part of the pandemic, we used a volunteer sample. To create weights for the observations to adjust for selection bias, we used the BRFSS as a reference survey for our target population. This section describes that adjustment.

From Figure 1 of the main paper, we see that there are several selection processes at work. First, individuals self-select to be in the volunteer pool, then self-select whether to answer the demographic survey, then of those invited from the volunteer pool by the quota sampling algorithm (Section 1.4), only 41% (=11382/27716) accepted the invitation, and only 29% (=8058/27716) were actually used in the final analysis. Although the distributions of the quota variables in the quota sample reflect those in the US population (by design), there are selection biases in other variables. For example, the education level for individuals in the final analysis data was much higher than that in the US population (see Table 2 of the main paper). We adjust for this imperfect representation due to the selection processes using the BRFSS survey as the reference.

The adjustment is conducted by weighting the observations by the pseudo-weights for each individual in the quota sample to denote the number of individuals in the target population that the individual represents. Three steps are involved to generate the final set of pseudo-weights.

Step 1 involves propensity model construction and estimation. The propensity model estimates the probability that each participant in the quota sample is selected from the target population. We start the model building by creating 16 variables created from the BRFSS questions that were also asked on the survey for this study (see Statistical Table 3). Note that the NCHS urban/rural variable that aligns with the urban/rural variable in the BRFSS differs from the urban/rural quota variable from the census. The race variable of 6 categories was collapsed into three categories (white only, black only and lower prevalence groups) to avoid sparse categories. Because we want the propensity model to ideally balance the marginal and higher order of joint distributions of covariates, we fit the model with all 16 covariates and all pairwise interactions. Unfortunately, that model produced highly variable estimated propensities with only 8058 observations. Next, we tried to use a model selection method to reduce the number of parameters (i.e., interactions terms) in the large model by applying backward selection. First, we removed all interaction terms with associated p-values for the effect > 0.25. This left 67 remaining interaction terms. Then iteratively, we remove the term with the largest p-value and re-fit the propensity model using the svyglm() R function in the survey R package (Lumley, 2020). We continue to iterate until all p-values of the remaining interaction terms are < 0.1. All the while, we keep all the main effect terms in the propensity model. The fitting of the propensity model used the observations in BRFSS and our survey, from which the estimated propensities were obtained for each individual in our study.

In Step 2, given the estimated propensities, kernel smoothed pseudo-weights were constructed (denoted by KW) (Wang, et al. 2020a). The KW method using the estimated propensities, as other propensity-based weighting or matching methods, makes the following assumptions. First, the reference survey sample (in our case the BRFSS), through weighting, properly represents the target population of interest. As desired, after adjustment the variables in Statistical Table 4 show similar distributions between BRFSS and Census. Second, all finite population units have a positive participation rate (i.e., each individual in the population has a positive propensity to volunteer to participate in the study). Third, we have conditional exchangeability with no unmeasured confounders. The third condition means that the probability for each individual in the population to participate in the study is not related to his/her seropositivity, after adjusting for measured variables (i.e., confounders) i.e., the 16 variables from BRFSS in Statistical Table 3. The KW method uses the estimated propensities as the measure of similarity, and fractionally distributes survey representative weights to the quota sample units based on a kernel smoothed distance in the estimated propensities. The choice of the KW method among various propensity-based weighting or matching methods was driven by its robustness to the propensity model misspecification while avoiding the very small, estimated propensities, which resulted in very large pseudo-weights and caused high variability in pseudo-weighted prevalence estimates. Wang et al. (2020a) showed the KW method when compared with its alternatives (i.e. Valliant and Dever 2011; Wang, et al, 2020b) tends to obtain estimates with smallest mean squared error.

Finally, in step 3, the kernel smoothed pseudo-weights were post-stratified on five quota variables (age, sex, race, Hispanics, and region) used in the propensity model to construct the final set of pseudo-weights. Poststratification adjustment is very common in practice, playing an important role in many national surveys, to increase precision of survey estimates (e.g. Valliant 1993). The poststratification cells defined by the five quota variables cover almost all of the census population (99.3%). The final poststratification-adjusted kernel smoothed pseudo-weight (psKW) is computed for each individual in the analytic data set.

The seropositivity prevalence in the entire survey population (i.e., undiagnosed adults in the US within the 51 state/districts) or subpopulations (e.g., individuals 70 and older) are estimated by psKW-weighted proportion. The standard errors of estimated seropositive rates were estimated by the Taylor linearization method, accounting for three sources of variabilities, i.e. the propensities estimated in step 1, differential KW pseudo-weights constructed in step 2, and poststratification adjustment in step 3 (Wang et al. 2020; Valliant 1993).

#### 1.6 Adjustment for Sensitivity and Specificity

We make adjustments for sensitivity and specificity for the serology assay.

To estimate specificity, we use a sample of negative controls ($n$ = 300) that was selected from a collection of frozen samples all collected before the start of the COVID-19 outbreak (serum collection date prior to January 2019). We assume the sample is approximately a simple random sample from the set of the negative samples from the US at the time that the data are collected. This could be a problem if our negative control sample had an unusually low or high rate of infection with another coronavirus that has cross-reactivity with the SARS-CoV-2 antibody assays that we use. This is unlikely to be a large problem because there does not appear to be strong cross-reactivity (see Figure 6, Klumpp-Thomas and Kalish et al, 2020). Furthermore, we do see similar reactivity to both spike variants, 614D and 614G (see Klumpp-Thomas et al, Jour of Infect Diseases, 2020). However, if there is a new coronavirus that is circulating during the study collection time, that was not circulating during the time the data for Figure 6 was collected, and that new coronavirus has cross-reactivity, then this will bias the results to have a higher seroprevalence estimator.

To estimate sensitivity, we use set of positive controls of $n$ = 56 people that are virologically-confirmed cases of COVID-19. For convenience, we make the assumption that the sample is approximately a simple random sample from the set of people who got infected with SARS-CoV-2, and that the distribution of the antibodies is similar between this sample and that of the undiagnosed sample for the study. There may be slight biases due to the positive controls being symptomatic, although the biases could go in either direction. If the antibody level of our positive control sample is biased upward (i.e., larger levels in the symptomatic positive controls than in the population of infected but undiagnosed with SARS-CoV-2), then our sensitivity estimate is likely higher than it really is, and hence our prevalence estimate will be biased lower (i.e., be an under-estimate). Conversely, the symptomatic individuals in our positive control sample could be symptomatic because their immune system was under-performing and hence had antibody levels that were lower than the ideal sample, and hence our prevalence estimate will be biased higher (i.e., be an over-estimate).

For any assay used to measure the prevalence, the standard adjustment that modifies the apparent prevalence estimate ($\widehat{AP}$) from the assay that accounts for the sensitivity estimate ($\widehat{Se}$) and the specificity estimate ($\widehat{Sp}$) gives an adjusted prevalence estimate ($\hat{P}$) as

$$\hat{P} = \frac{\widehat{AP} + \widehat{Sp} - 1}{\widehat{Se} + \widehat{Sp} - 1},$$

(see, e.g., Rogan and Gladen, 1978). To calculate confidence intervals for the adjusted prevalence, we use a slight generalization of the melding method, that uses lower and upper confidence distributions on functions of independent estimators (Fay, et al, 2015). This allows us to create a confidence interval for the prevalence that accounts for the variability in $\widehat{AP}, \widehat{Sp},$ and $\widehat{Se}$ without relying on asymptotic normality. For $\widehat{Sp}$ and $\widehat{Se}$ we use the lower and upper confidence distributions associated with the exact binomial confidence interval for each. For example, for a binomial random variable, if we observe x out of n, then the lower confidence distribution for the binomial parameter is Beta(x,n-x+1) and the upper confidence distribution is Beta(x+1,n-x). For $\widehat{AP},$ we use the method of Korn and Graubard (1998) to derive the lower and upper confidence distributions, which gives a lower confidence distribution of Beta(x*,n*-x*+1) and an upper confidence distribution of Beta(x*+1,n*-x*), where n*= $\widehat{AP}(1-\widehat{AP})/s^2$, where s is the standard error of $\widehat{AP}$ described at the end of Section 3.5, and x*=$n^* \times \widehat{AP}$. We use the melding method to get the confidence interval.

Let $B_{se}^L, B_{se}^U, B_{sp}^L, B_{sp}^U, B_{ap}^L, and\ B_{ap}^U$ be the lower and upper confidence distribution random variables associated with the three estimators, and let q(a,W) be the a[th] quantile of a random variable W, then the 95% confidence interval is

$$\left\{q\left(0.025, \frac{B_{ap}^L+B_{sp}^L-1}{B_{se}^U+B_{sp}^L-1}\right), q\left(0.975, \frac{B_{ap}^U+B_{sp}^U-1}{B_{se}^L+B_{sp}^U-1}\right)\right\},$$

which is estimated by Monte Carlo simulation $10^5$ replications. Lower limits less than 0 are replaced with 0.

#### 1.7 Applications of the Methods for Specific Statistics

The previous sections have described the methods for our main estimators for the total population and subpopulations. Here we describe additional statistics calculated.

##### 1.7.1 Analysis of Recombinant Antibody Dilution Curves and 4PL

A dilution curve of 1 mg/ml recombinant anti-SARS-CoV-2 S1 RBD monoclonal IgG, IgM and IgA antibody (GenScript) was generated in sample loading buffer then run utilizing our standard ELISA protocol. The resulting OD values were then plotted against recombinant antibody concentration in loading buffer to generate a dataset suitable for fitting of a sigmoidal model to generate a concentration value that could be translated to compare multiple serologic assays and serosurveys pending the other assay was also run against this recombinant curve. Sigmoidal four parameter logistic models were fit using GraphPad Prism v9. The results of these interpolations are available in Table S2.

##### 1.7.2 Estimation of Ratio of Counts of Undiagnosed and Seropositive Over Diagnosed

We describe the estimate of the ratio of the number of "undiagnosed cases" (US adults previously undiagnosed with SARS-CoV-2 infection by PCR, that are seropositive for SARS-CoV-2 antibodies) over "diagnosed cases" (US adults previously diagnosed by PCR) during the time period of the study. Our main estimate of the paper is the rate of undiagnosed cases per total undiagnosed US adults over the study collection time. Loosely speaking, our estimate of the ratio for US adults uses

$$\frac{\text{(number previously undiagnosed)} \times \text{(rate of undiagnosed cases per undiagnosed)}}{\text{(number previously diagnosed)}}$$

What makes this estimator difficult is that the data are collected over a few months during which the numbers of cases diagnosed are changing dramatically. We describe the data used to estimate the number previously diagnosed and undiagnosed cases at any specific date in two separate sections. Then we describe how the time element is used to obtain our final estimate.

We used The COVID Tracking project (covidtracking.com), and code derived from Chow et al (2020), to get, for any date of the study, the total cumulative number of virologically-confirmed cases in the US. To estimate that number for adults, we multiplied the totals for each date by 0.959, which is the ratio of adult virologically-confirmed cases on September 19, 2020 (estimated as the total cumulative cases on that date minus the estimated amount that were in children 5-17 [Leeb, et al, 2020], omitting children under 5) over the total cumulative cases on that same date.

To estimate the number of adults previously undiagnosed for any date of the study, we can just subtract the number of previously diagnosed cases by that date from the total number of adults in the US at that date. To estimate the number of adults in the US at date t, we start with the total number of people in the US at date t based on the US census population clock (US Census, 2020, U.S. and World Population Clock), and multiply by the proportion of the population that are adults, about 0.7752 (estimated from 2018 census data used for the quota sampling).

To precisely define our estimator, we introduce notation. Let $R_{udc/ud} = \sum_{i=1}^{n} s_i\, y_i,$ be our weighted estimator of the rate of previously undiagnosed cases who were found to be seropositive per all undiagnosed individuals, where $s_i$ and $y_i$ are, respectively, the standardized weights (standardized to sum to 1), and the binary seroprevalence results for the $i^{th}$ individual in the final data set. Let $t_1,\ldots,t_n$ be the sample collection dates. Let $P_t(t)$ and $N_{dc}(t)$ be the estimates of the total population of US adults and the previously diagnosed US adults at date t.

Our estimate of the ratio of the number of previously undiagnosed seropositive US adults over the number of previously **diagnosed** US adults is:

$\frac{\{\overline{P}_t - \overline{N}_{dc}\} R_{udc/ud}}{\overline{N}_{dc}} = 105.06 \times R_{udc/ud},$ where $\overline{P}_t = \frac{1}{n}\sum_{i=1}^{n} P_t\,(t_i)$ and $\overline{N}_{dc} = \frac{1}{n}\sum_{i=1}^{n} N_{dc}\,(t_i)$.

To get the 95% confidence interval, we multiply the lower and upper limits for $R_{udc/ud}$ by 105.06. As a sensitivity analysis, if we use the weighted means, $\overline{P}_t^{w} = \sum_{i=1}^{n} s_i\, P_t\,(t_i)$ and $\overline{N}_{dc}^{w} = \sum_{i=1}^{n} s_i N_{dc}\,(t_i),$ in place of the unweighted means the multiplicative factor is nearly the same, 104.08.
